# Enhancing the Human Health Status Prediction: the ATHLOS Project

**DOI:** 10.1101/2021.01.19.21250076

**Authors:** Panagiotis Anagnostou, Sotiris Tasoulis, Aristidis G. Vrahatis, Spiros Georgakopoulos, Matthew Prina, José Luis Ayuso-Mateos, Jerome Bickenbach, Ivet Bayes, Francisco Félix Caballero, Laia Egea-Cortés, Esther García-Esquinas, Matilde Leonardi, Sergei Scherbov, Abdonas Tamosiunas, Aleksander Galas, Josep Maria Haro, Albert Sanchez-Niubo, Vassilis Plagianakos, Demosthenes Panagiotakos

## Abstract

Preventive healthcare is a crucial pillar of health as it contributes to staying healthy and having immediate treatment when needed. Mining knowledge from longitudinal studies has the potential to significantly contribute to the improvement of preventive healthcare. Unfortunately, data originated from such studies are characterized by high complexity, huge volume and a plethora of missing values. Machine Learning, Data Mining and Data Imputation models are utilized as part of solving the aforementioned challenges, respectively. Towards this direction, we focus on the development of a complete methodology for the ATHLOS (Ageing Trajectories of Health: Longitudinal Opportunities and Synergies) Project - funded by the European Union’s Horizon 2020 Research and Innovation Program, which aims to achieve a better interpretation of the impact of aging on health. The inherent complexity of the provided dataset lie in the fact that the project includes 15 independent European and international longitudinal studies of aging. In this work, we particularly focus on the HealthStatus (HS) score, an index that estimates the human status of health, aiming to examine the effect of various data imputation models to the prediction power of classification and regression models. Our results are promising, indicating the critical importance of data imputation in enhancing preventive medicine’s crucial role.

## 1 Introduction

We live in the “Big Data” era, which offers a great potential for revolutionizing all aspects of society, including the healthcare domain [1]. The recent advancements in medical and health domains generate, with an exponentially increasing rate, several heterogeneous data types, such as medical, demographic, molecular, cellular, and so on, offering great potential for personalized healthcare [2]. Digitization in medicine and healthcare has revolutionized how various biological processes and complex diseases are interpreted, since there is the possibility for analysis and knowledge extraction from large scale data [3]. In this direction, longitudinal studies, such as cohort studies and natural experiments, offer a platform to study more profound challenges in the biomedical and healthcare domain [4]. The predominant way to deal with the inherent data complexity is the Machine Learning frameworks [5], which allow the exploration, development and construction of algorithms that can learn from data and produce accurate predictions [6, 7].

Meanwhile, clinical and population health research usually includes questionnaires within longitudinal studies, creating a significant challenge regarding the missing values for several apparent reasons (non-response, loss tracking, etc.) [8]. The proper way to deal with this critical limitation is to impute the missing information without losing the imposed data structure [9]. The recent literature is plentiful with imputation methods for longitudinal studies [9–14], while it has been shown that such a pre-process step can lead to improvements in various Machine Learning tasks, such as the data classification [15]. Longitudinal studies often contain categorical information, which is restricted-transition variables. This information is usually incomplete; hence its imputation is a significant challenge [9]. The difficulty lies in the fact that there is little guidance on whether these restrictions need to be accommodated when using imputation methods [16]. Nevertheless, most researches on this aspect focus on continuous data with fewer recommendations for categorical data [17].

A further essential challenge when dealing with longitudinal studies is the accuracy and validity of prediction models [18]. Towards this perspective, we study the prediction power in classification and regression tasks utilizing various imputation methodologies. Our aim is to enhance the predictability of machine learning tasks on a large-scale longitudinal study, characterized by high rate of missing values, and simultaneously mine new insights and knowledge for health status across the human age. For this purpose, we employ a harmonized dataset produced within the ATHLOS (Ageing Trajectories of Health: Longitudinal Opportunities and Synergies, http://athlosproject.eu/) Project (EU HORIZON2020–PHC-635316), which integrates 15 independent European and international longitudinal studies of aging.

The ATHLOS Project is based on improving a critical factor in the healthcare domain, extending the healthy lifespan. Its importance lies in the fact that in the last decades is observed a remarkable worldwide population aging due to the increase in life expectancy [19]. The healthcare demand increases along with the increasing number of elderly populations, which consists of a high cost every year for states [20]. Health status measurements are an immensely important indicator that helps prevent human health near or even in the distant future [21]. Under this perspective, the ATHLOS project aims to better study the impact of aging on health by developing a new single measure of health status. The project intends to identify patterns of healthy aging trajectories and their determinants, the critical points in time when changes in trajectories are produced, and propose timely clinical and public health interventions to optimize and promote healthy aging.

Furthermore, one of the unique features of ATHLOS is a metric of health, called Health-Status (HS), proposed in [22]. It is a novel index for human health condition calculation using an Item Response Theory (IRT) approach. This measure is based on specific characteristics that are part of the ATHLOS features list and has shown its reliability in several studies [23–38]. The challenge that now emerges is to discover meaningful relationships between the wide range of features found across various HS studies. Efficiently predicting HS through features that have been considered unrelated or not informative and thus excluded from its designing process would promote its expandability in further health-related studies while opening up new avenues for HS optimization.

This study focuses on strengthening the prediction power of the ATHLOS HealthStatus score in terms of classification and regression through various imputation methods. A large variety of both well-established and cutting-edge imputation and prediction methodologies were applied. More specifically, we applied five and six imputation and prediction methods, respectively by covering the broader range of imputation and prediction methods family. Our study shows that applying different imputation methods can greatly affect the various classification and regression schemes in terms of their accuracy and robustness. By discovering the most appropriate and methodological framework, we attempt to offer new insights and identify a better interpretation of the impact of aging on health status. The aforementioned outcomes can be considered promising, completing the puzzle of the strong outcomes that have emerged from previous studies of ATHLOS [23–38].

## 2 Methods & Experiments

Five imputation methods were applied (Mean [39], Linear Regression [40], Dual Imputation Model [41], Multiple Linear regression [42], and Vtreat method [43]) the harmonized ATHLOS dataset. Subsequently, the imputed matrices have been used as input to the six mentioned traditional and state-of-the-art regression and classification tools (Logistic/Linear Regression [44], k Nearest Neighbors [45], Random Forests [46], Extreme Gradient Boosting (XGBoost) [47], and two Deep Neural Network models). Details about the methodological pipeline, the brief analysis of all algorithms and the experimental setup are given in the following subsections.

### 2.1 Data preprocessing - Cleaning and Preparation

The ATHLOS project provides a harmonized dataset [23], built upon several longitudinal studies, originated from five continents. More specifically, it contains samples coming from more than 355,000 individuals who participated in 17 general population longitudinal studies in 38 countries. In this paper, we used 15 of these studies, namely the 10/66 Dementia Research Group Population-Based Cohort Study [24], the Australian Longitudinal Study of Aging (ALSA) [25], the Collaborative Research on Ageing in Europe (COURAGE) [26], the ELSA [27], the study on Cardiovascular Health, Nutrition and Frailty in Older Adults in Spain (ENRICA) [28], the Health, Alcohol and Psychosocial factors in Eastern Europe Study (HAPIEE) [29], the Health 2000/2011 Survey [30], the HRS [31], the JSTAR [32], the KLOSA [33], the MHAS ([34]), the SAGE [35], SHARE [36], the Irish Longitudinal Study of Ageing (TILDA) [37] and the the Longitudinal Aging Study in India (LASI) [38].

The 15 general population longitudinal studies utilised in this work consist of 990,000 samples in total, characterized by 184 variables, of witch two are considered as response variables and the rest as independent variables. Response variables are the raw and the scaled HealthStatus scores for each individual. Regarding the independent variables (please see supplementary material, sheet S1), nine variables were removed including various indexes (sheet S2), 13 variables were removed including obviously depended variables that cannot be taken into account (sheet S3), and six variables were removed including information that cannot be considered within a prediction model (sheet S4).

It is worth mentioning that the 47 variables (sheet S5) that were originally used to calculate the HS score [22] have been excluded. These features create a statistical bias regarding the HS, which is the response variable in our analysis. Keep in mind, that these variables have been previously identified as more critical in defining the HS, as such we expect that using the rest for predicting it would be a challenging task, which however may reveal new insights. Finally, removing the samples for which the HS metric is missing, the resulting data matrix is constituted by 770,764 samples and 107 variables.

### 2.2 Missing Data Imputation

A brief analysis of all imputation tools used (Mean Imputation, Linear Regression, Dual Imputation Model, traditional Multiple Imputations, Vtreat) is provided. Mean Imputation (MI), Linear Regression (LR) belongs to the single imputation methods family having the procedure to replace an unknown missing value by a single value. MI is a simple process based on the calculations of the mean value of the non-missing values for each response variable with a numerical data type. Missing values are filled with the respective mean value for each response variable. It is a relatively fast and straightforward approach, giving good results for relatively small numerical datasets. However, it exhibits various limitations, such as it does not consider the correlation among the response variables along with the data uncertainty in the imputations [48]. In the Linear Regression (LR) imputation, the missing values are replaced with predicted values that arise from applying a linear regression model in the non-missing data. The idea behind this process is the fact that the variables tend to be correlated. Hence, the predicting values should be affected by the observed data [40].

On the other hand, the Dual Imputation Model (DIM) and the Multiple Linear Regression Imputation (MLR) belong to the MI family where the missing values are replaced by *n* simulated values (*n >* 1) while its main assumption is that data are missing at random. The MI frameworks deal with the uncertainty about which values to impute, a major limitation of singular imputation methods. MLR imputation reduces this limitation by combining several different plausible imputed datasets. Initially, multiply imputed datasets are constructed and then standard statistical methods are applied to fit each one. Under the MI perspective, a recently proposed imputation method is the Dual Imputation Model (DIM) [41], which approximates an offset between the data distribution of the observed and missing data. This approximation is performed by iterating over the response and imputation models, assuming that the response as the imputation model has the same predictors.

Vtreat [43] is a recent imputation method that differs from similar traditional tools since it considers the uncertainty in the imputations. It includes a unique strategy with respect to the type of the response variable, creating additional variables along with the well-known dummy variables process. For a numerical type response variable, one more dummy variable is created to address the high cardinality categorical variables. Two dummy variables are created to cope with the novel (or rare) levels found in each categorical variable of the data set. For a categorical type response variable, one more dummy variable is created to address the high cardinality categorical variables. One dummy variable is created to cope with the novel (or rare) levels found in each categorical variable of the dataset. This inherent feature enhances the knowledge discovery of ultra-noisy data. Furthermore, vtreat is useful in the modeling process because it allows the model to re-estimate the response on systematically missing values. This property motivated us to focus our attention upon it since the nature of longitudinal studies imposes the emergence of systematically missing values at a high degree.

Regarding the experimental study and for the MI method, 141 dummy variables were created during the conversion of 66 categorical predictor variables. Mean values from each of the 34 remaining predictor variables values were calculated by replacing the respective missing values indexes. For the LR Imputation, 141 dummy variables were constructed following the same process for creating the dummy variables. A simple model was created for the remaining 34 predictor variables:

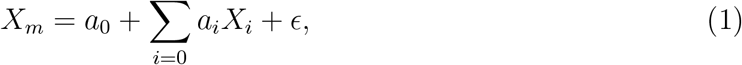

where *X*_*m*_ is the predictor variable to be imputed, *X*_*i*_ is the list of *n* complete predictor variables, *a*_*i*_ are the regression coefficients, *a*_0_ is the intercept, and is *ϵ* the normally distributed error.

Regarding the application of MLR (Multiple Linear Regression) imputation, we followed the multiple imputation main steps by using the Linear Regression model to perform the *m* complete data analyses. Initially it defines the posterior predictive density *p*(*Y*_*mis*_ | *X, R*), where *Y*_*mis*_ indicates the missing parts of *Y, X* is the 34 predictor variables, and *R* is the 770, 764 × 33 binary matrix indicating the elements of *Y* that are observed (*R*_*ij*_ = 1 if *Y*_*ij*_ is observed). Note that *Y* indicates the partially observed outcome variables in a sample of size *n* = 770, 764. Subsequently, imputations are generated based on this density by concatenating *m* = 5 complete datasets. In the next step, a Linear Regression model was applied on each completed data matrix performing *m* = 5 complete data analyses, while an ensemble criterion exports the outcome for each point. Here, we employ the R implementation of the MICE (Multivariate Imputation by Chained Equations) [49] for the MLR model.

In the DIM (Dual Imputation Model), we integrated the multiple imputation along with the doubly robust weight frameworks to deal with misspecification included on ATHLOS data with missing at random values. Briefly, let the observed data be **V** = (**Y**_obs_, **R, X, U**), and *P* (***θ, a*** | **V**) ∝ *P* (**V** | ***θ, a***)*p*(***θ, a***), and *p*(***θ, α***) is the prior density. The main equation of DIM which defines the posterior predictive distribution of missing data given the observed data, is:

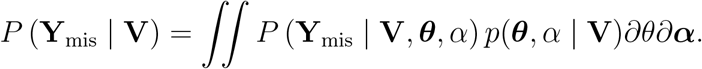

Initially, we imputed the missing values by a random process taking into account the observed ATHLOS data. Then an iterative process was applied by (i) drawing a random value 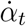 from its posterior distribution, (ii) calculating the propensity scores 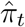 given the drawn value 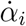, (iii) adding an additional predictor into the imputation model, (iv) estimating the parameters of the imputation model for the observed values, (v) drawing a random value from their posterior distributions, and (vi) imputing the missing value adding a noise factor. We implemented this iterative process ten times, as suggested by [41].

The cutting-edge vtreat was applied on ATHLOS dataset following its previously described principles. We imputed the missing values through 458 dummy variables created for the continuous numerical response variable and 392 dummy variables created for the categorical response variable. The resulting matrices are given as input in the next step of the proposed framework, the significance pruning process. Each variable was evaluated based on its correlation with the HealthStatus (HS) [22] score (response variable) in order to isolate the most significant variables. More specifically, we applied a treatment plan by decoding and removing the noisy variables. For this purpose, we calculated their correlation with HS using the F-test, since it is a numeric variable and a *X*^2^-Test for the categorical target created with respect to the classification tasks.

The significance of each predictor variable was calculated using F-statistic on the single-variable regression model and *X*^2^-statistic on the logistic model between the response variable and each predictor. Subsequently, we pruned the variables with higher significance value than the threshold 1*/n*_*vars* in both cases, where *n*_*vars* indicates the resulting number of variables obtained from the matrix imputation step. As a result of the significance pruning process, for the numerical target, 51 noisy variables were removed. The rest 407 variables were normalized using the linear regression model. Thus, for each predictor variable we have:

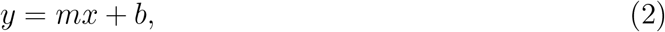

where *x* is a predictor variable, *m* is the linear regression coefficient, and *b* is the intercept. Then, scaling and centering takes place with the following procedure:

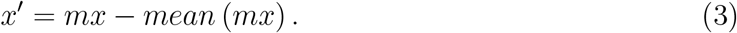

For the categorical response variable, about 51 noisy variables where removed, resulting in 351 remaining variables. Subsequently, the predictor variables were normalized using the logistic regression model for each predictor variable:

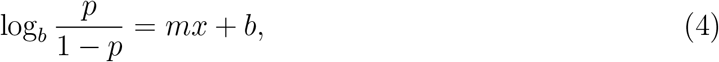

where *p* = *P* [*y* = *TRUE*] and *y* is the response variable, *x* is a predictor variable, *m* is the logistic regression coefficient and *b* is the intercept. Similarly, scaling and centering of each predictor variable took place with the following procedure:

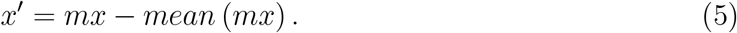

It is worth mentioning that all five methods utilize the dummy variable creation process for the categorical variables. In this approach, each categorical level is represented separately, including the values 0 and 1 for the participation or not in a specific level. The level that is not represented contains zeros on all dummy variables, acting as the reference level [50]. In our workflow, we deal with missing values of all categorical variables following the principles highlighted in [51]. The missing values are considered as regular categorical levels and are included in the dummy variable process. Concluding, five independent imputed datasets were generated and were used as input in the next section to examine the prediction power from various Machine Learning algorithms.

### 2.3 Prediction Methods

Six different classification and regression models, namely Logistic/Linear Regression, kNN, Random Forests, XGBoost, and two Deep Neural Network (DNN) models, have been applied on the five resulting datasets retrieved by applying the five imputation methods. Briefly, let *A* ∈ *R*^*s*×*d*^ be the ATHLOS dataset, where *s* is the number of samples, and *d* is the number of predictor variables. Let *H* ∈ *R*^*s*×1^ denote the response variable, a vector with the previously described HealthStatus (HS) score. For the classification process, a three-class categorization was considered after consulting previously similar categorization [52]. More precisely, the three groups are defined based on HS scores as follows: scores lower than or equal to 30, scores higher than 30 and lower than or equal to 50, and scores higher than 50. Linear and Multinomial Logistic regression was used for the regression and classification tasks, respectively. In linear regression, we used HS as the continuous dependent variable, while in logistic regression we used the aforementioned three-class categorization of HS. We applied kNN classification, searching for 5 nearest neighbors to assign each sample to one of the three classes using the Euclidean distance. In kNN regression, we calculated the average of the HS values of the k-Nearest Neighbors search of a given test point. Let *H* ∈ *R*^*s*×1^ denote the set of *s* training points that contain the HS values. The kNN estimator is defined as the mean function value of the nearest neighbors [53, 54]. The Random Forests (RF) regression operates in a similar way, with the main difference being that the final prediction output is the average of all trees’ output. RF method builds N trees, using randomization to decrease the correlation between them and achieve high accuracy. In this respect, trees operate on a random subset of the data. Furthermore, for each tree node, a random subset of the variables is considered. In our analysis, we use 100 trees and the *M*_*try*_ variable was defined as *p/*3 and 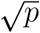, where *p* are the number of variables. Extreme Gradient Boosting (XGBoost) is a novel classifier based on an ensemble of classification and regression trees [47]. Let the output of a tree be:

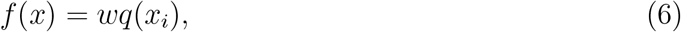

where *x* is the input vector and *wq* is the score of the corresponding leaf *q*. The output of an ensemble of K trees will be:

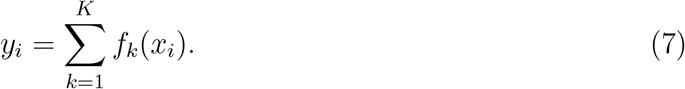

The XGBoost algorithm tries to minimize the following objective function *J* at step *t*:

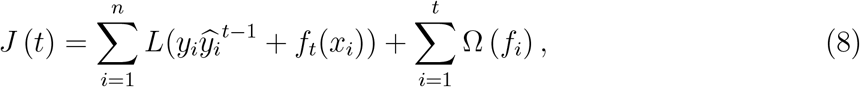

where the first term contains the train loss function *L* (e.g. mean squared error) between real class *y* and output *ŷ* for the n samples, and the second term is the regularization term that controls the complexity of the model and helps to avoid overfitting.

We also applied two Deep Learning architectures using the well-known Backpropagation (BP) method (based on the Gradient Descent method) for the training phase. BP requires a vector of input patterns *x* and the corresponding target *y*. Given an *x*_*i*_ where *i* corresponds to an individual input sample, the model produces an output *o*_*i*_. The main aim is to reduce the error in the training process through these proper weights that minimize the following cost function:

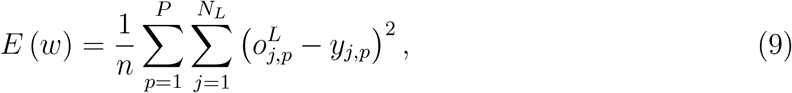

where *P* the number of patterns, 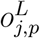 the output of *j* neuron of layer *L, N*_*L*_ is the number of neurons in output layer, *y*_*j,p*_ is the desirable target of *j* neuron of pattern *p*. The BP algorithm is utilized to minimize the cost function *E*(*w*), as mentioned above. The aim of optimization algorithm it to find the minimized 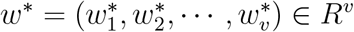, such that:

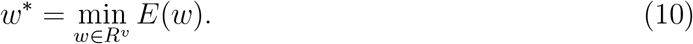

We selected the Deep Learning strategy to examine its behavior in avoiding underfitting or overfitting on ATHLOS training step since the examined imputation methods varying in terms of the number of features. More specifically, the first DNN (*DNN*_1_) consisted of two hidden layers of 100 neurons and one output layer of one neuron, while the second network (*DNN*_2_) has only one hidden layer with 100 neurons. In order to avoid the gradient vanishing problem, the ReLU activation function is utilized in hidden layers [55]. Since the response variable takes values in the range of (0, 100) to produce comparable results, the linear activation functions are utilized in the output layer. Finally, the learning rate is set to 0.001.

## 3 Results & Discussion

All executions were validated through the Monte Carlo cross-validation technique [56], creating multiple random splits of the dataset into train and test by selecting 100,000 and 10,000 samples, respectively. We implement 80 independent executions assuring that each sample was used as a test sample at least once. The regression performance of each imputation method was calculated using the R-squared measure [57] and the Root Mean Squared Error (RMSE) [58] upon the test set. Also, Accuracy, F1-score, Sensitivity, and Specificity were evaluated to measure the classification performance. The results are summarized in the Figure 1 and Table 1.

**Table 1:** Comparison of five imputation methods (Linear Regression (LR), Mean, Multiple Linear Regression (MLR), Dual Imputation Method (DIM), Vtreat) in regression tasks using 6 different regression techniques (Deep Neural Network (DNN) 1, DNN2, k-Nearest Neighbors (kNN), Linear Regression (LR), Random Forests (RF), XGBoost). The table contains the mean (standard error) values (%) of the R-squared measure, the mean (standard error) values of Root Mean Square Error (RMSE) as well, from 80 independent executions. The best value among imputation methods for each classifier is depicted in bold and the highest value of all imputation methods for all classifiers is depicted in bold italics.

|  | Linear Regression | kNN | Random Forests | XGBoost | DNN1 | DNN2 |
| --- | --- | --- | --- | --- | --- | --- |
| R-squared |  |  |  |  |  |  |
| LR | 53.06 (0.61) | 2.00 (0.29) | 55.20 (0.86) | 55.53 (1.44) | 9.83 (0.42) | 5.75 (0.36) |
| Mean | 53.30 (0.81) | 22.19 (0.72) | **55.91 (0.53)** | 56.66 (0.64) | 40.55 (0.56) | 19.70 (0.69) |
| MLR | 53.02 (1.04) | 2.55 (0.30) | 54.83 (1.28) | 56.21 (1.24) | 44.67 (0.67) | 0.01 (0.00) |
| DIM | 52.94 (0.83) | 1.85 (0.36) | 54.47 (0.79) | 55.72 (0.86) | 9.44 (0.37) | 3.69 (0.27) |
| Vtreat | **53.42 (0.63)** | **48.40 (1.10)** | 55.90 (0.70) | **57.84 (0.78)** | **55.84 (0.63)** | **45.04 (86.00)** |
| Root Mean Square Error |  |  |  |  |  |  |
| LR | 6.841 (0.048) | 10.418 (0.110) | 6.770 (0.104) | 6.658 (0.080) | 9.448 (0.134) | 9.648 (0.137) |
| Mean | 6.822 (0.037) | 9.003 (0.062) | **6.642 (0.037)** | 6.579 (0.041) | 7.579 (0.089) | 8.801 (0.095) |
| MLR | 6.841 (0.078) | 10.363 (0.095) | 6.743 (0.054) | 6.641 (0.044) | 9.473 (0.126) | 9.978 (0.114) |
| DIM | 6.870 (0.070) | 10.484 (0.101) | 6.728 (0.046) | 6.640 (0.057) | 8.824 (0.115) | 9.787 (0.127) |
| Vtreat | **6.808 (0.067)** | **7.279 (0.078)** | 6.659 (0.054) | **6.573 (0.048)** | **6.653 (0.090)** | **7.299 (0.097)** |

**Figure 1:**
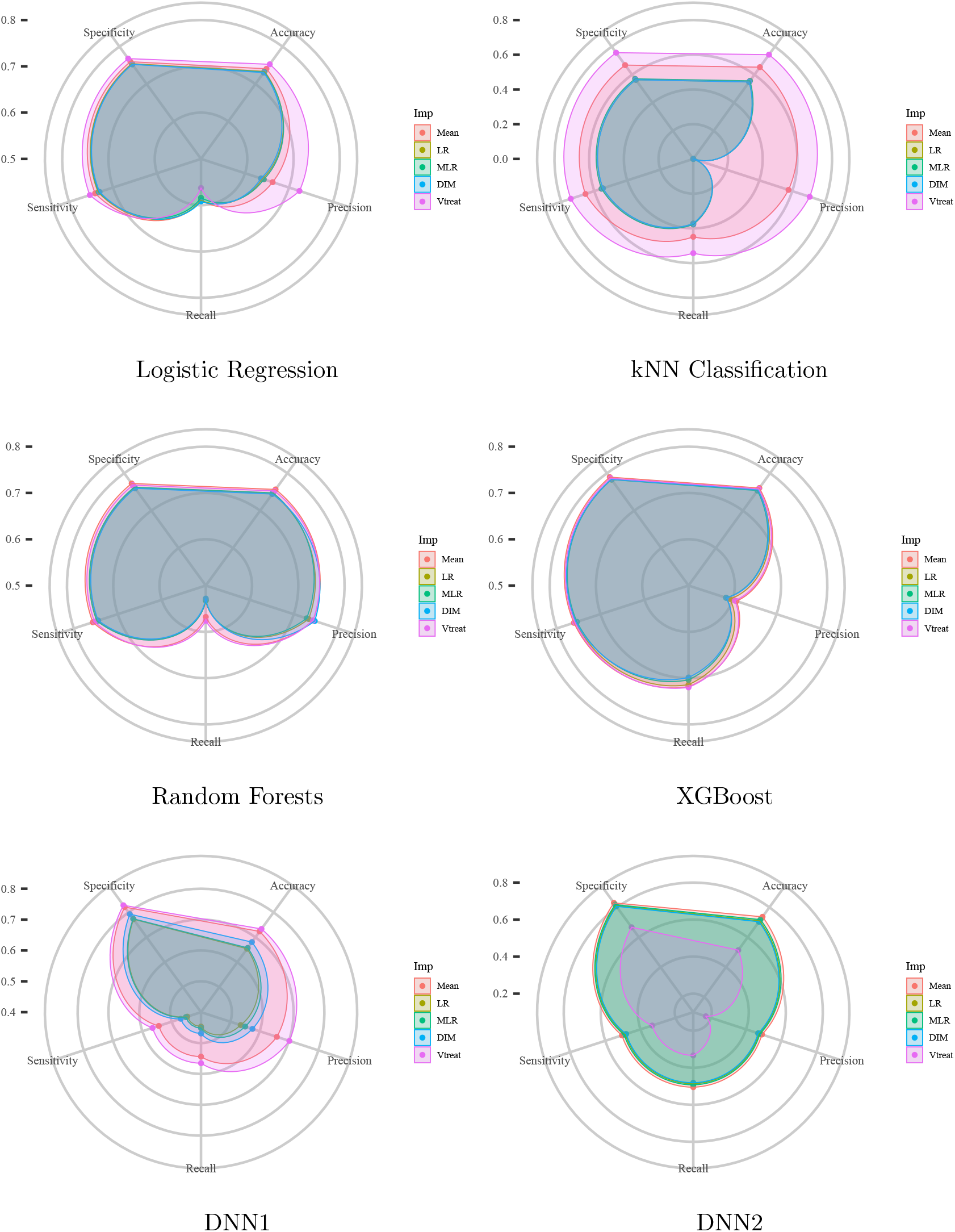
Each radar plot contains the visual representation of the classification results for each imputation method used in this paper. The methods are: Mean (Mean) imputation, Linear Regression (LR) imputation, Multi Linear Regression (MLR) imputation, Dual Imputation Model (DIM), and Vtreat imputation. The axes of the radar plots are metrics accuracy, precision, recall, sensitivity, and specificity. Finally, there is one radar plot for each of the classification models utilized. Namely, for the implementation of the Logistic Regression model, the kNN Classification model, the Random Forests model, the XGBoost model, and the two Deep Neural Network models (DNN1 and DNN2).

In general, we observed that the Vtreat imputation method consistently improves both the classification and regression performance compared to the other methods. Its superiority appears in almost all evaluation measures and cases. Interestingly, we observe a significant improvement for the deep neural network and k-Nearest Neighbors concerning classification and regression. A minor difference is observed in logistic and linear regression for classification and regression performance, respectively. Additionally, we observe that the combination of XGBoost and Vtreat outperforms any other method/combination for both regression and classification tasks. Given that XGBoost is an efficient, cutting-edge classification technique, Vtreat reveals its true potential by enhancing its performance. Apparently, the better structure offered affects tree-based ensemble algorithms known to be appropriate for structured data. However, we also observe that Vtreat significantly improves the performance of both DNN models, known to perform better for unstructured data, such as images and text, further showing its potential in the data imputation task.

### 3.1 Visualizing the prediction power

Visualization is a central part of ML approaches, since it offers insights into the data structure. Most visualization approaches apply a dimensionality reduction technique to transform the multidimensional data into two or three dimensions, while preserving their relationships. Here, we focus on visualizing each imputation method’s predictive power through a two-dimensional representation. For this purpose, we employed the Principal Components Regression (PCR) technique to predict the Healthstatus score. Through this, we can investigate prediction power along with the corresponding two-dimensional representation produced by the respective Principal Component Analysis (PCA) method. Our motivation also lies in PCR’s advantages in both statistical and computational aspects, which reduce the variance estimation at the expense of other bias and offers a better model fitting respectively [59].

Briefly, let *A* ∈ *R*^*s*×*d*^ be the ATHLOS dataset, where *s* is the number of samples, and *d* is the number of predictor variables after applying the significant pruning step. Let *H* ∈ *R*^*s*×1^ denote the vector of HealthStatus scores. Through PCR we initially perform PCA on the centered data matrix *A* and subsequently only consider the first two principal components, producing the matrix *A*^*PC*^ ∈ *R*^*s*×2^. For the *N* number of principal components, the formula for the HS evaluation is calculated as:

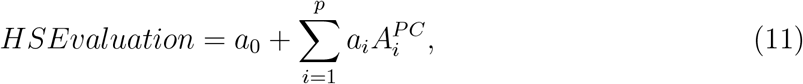

where *a*_0_ is the intercept, *a*_*i*_ is the *i*^*th*^ regression coefficient and 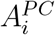 is the *i*^*th*^ principal component resulted from PCA. The PCR technique is applied upon all five datasets produced by the application of each corresponding imputation method.

Initially, we observe that the low, medium and high values of HealthStatus score appear better separability for the Vtreat method. In addition, observing the class distribution with respect to the first Principal Component, a clear distinction between classes is only achieved for the Vtreat method, while the respective distributions arising from LR, Mean, and DIM imputation methods have quite similar behavior (see Figure 2 for details).

**Figure 2:**
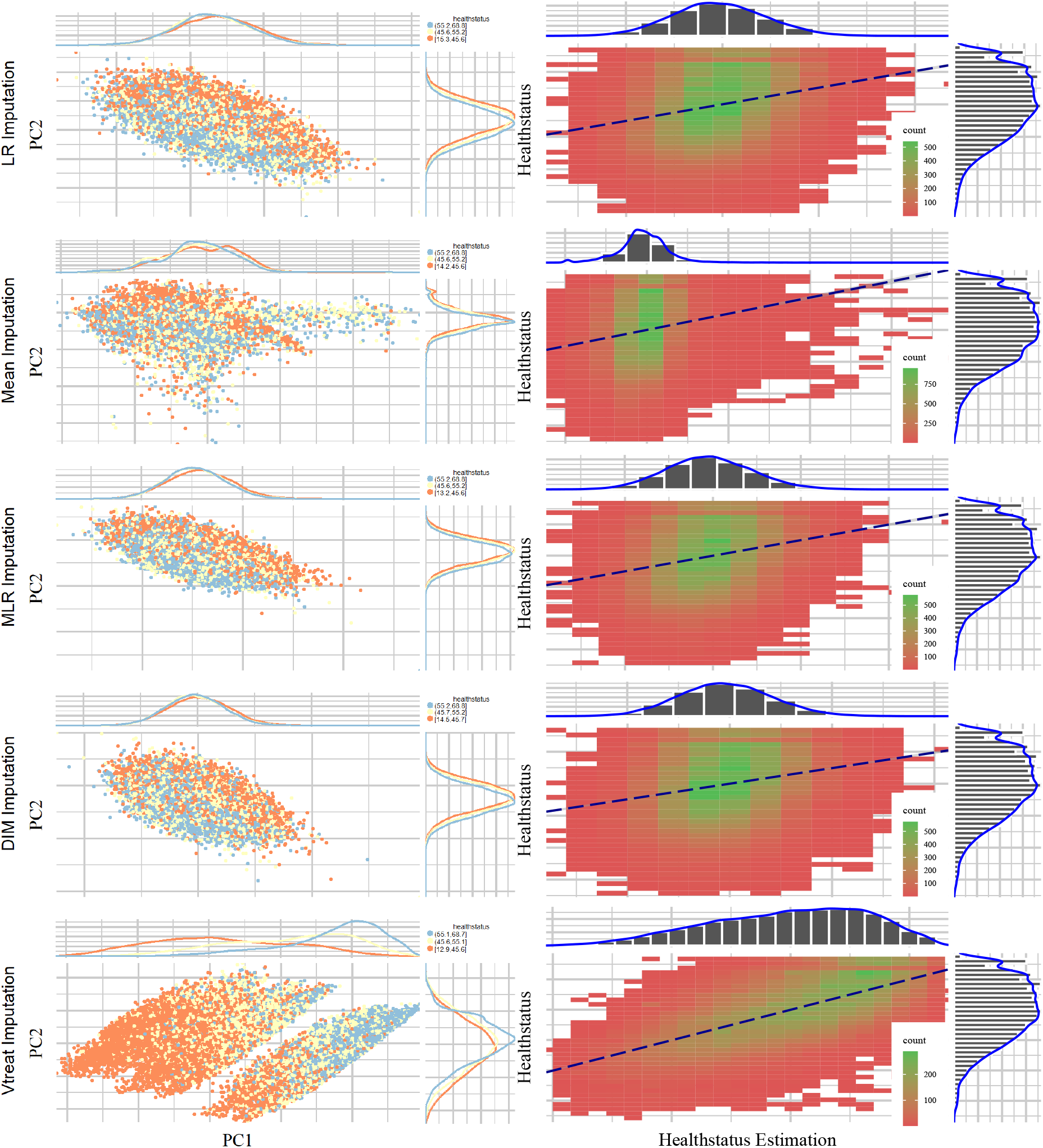
Scatter plots (left column) depict the first two principal components of PCA performed on the five imputed ATHLOS datasets using Linear Regression, Mean, Dual Imputation Model, and Vtreat imputation. Circular points with orange, yellow, and light blue colors illustrate the low, medium, and high HealthStatus scores. Above and right to each scatter plot, their data distribution is illustrated. Heatmap-Scatter plots (right column) depict the correlation of predicted and real HS score of the five imputation methods using the Principal Components Regression (PCR) technique. The red to green color graduation of boxes indicates the number of samples from low to high amounts, respectively. Above and right to each heatmap-scatter plot is illustrated the marginal distribution of the HS and the HS estimation as univariate histograms with a density curve on the vertical and horizontal axes of the scatter plot, respectively.

Utilizing the PCR technique to visualize regression efficiency (see Figure 2, right column) allow us to observer that the best performance is achieved by the Vtreat method, indicated by the higher correlation between predicted and real HS scores. Zooming into the respective heatmap-scatter, we can see the high number (green graduation in boxes) of samples across the dashed line, highlighting the optimal regression as reported in Table 2. We also observe all other methods perform poorly compared to Vtreat, while the mean imputation method behaves differently from the other two, with almost identical results.

**Table 2:** Comparison of 5 imputation methods (Linear Regression (LR), Mean, Multi Linear Regression (MLR), Dual Imputation Method (DIM), Vtreat) using the Principal Components Regression technique. The table contains the (%) of the R-squared measure, the mean (standard error) values of Root Mean Square Error (RMSE) as well. The best value among imputation methods for each measure is depicted in bold.

|  | LR | Mean | MLR | DIM | Vtreat |
| --- | --- | --- | --- | --- | --- |
| R-squared | 6.07 | 2.89 | 5.93 | 4.9 | **43.5** |
| RMSE | 9.705 | 9.862 | 9.733 | 9.767 | **7.545** |

### 3.2 Feature Importance for HealthStatus prediction

In this Section, our aim is to identify the factors that significantly affect HealthStatus prediction. Based on the indicated performance evaluation within the aforementioned experimental analysis, we captured this information by isolating the variable importance measure of the XGBoost algorithm when applied to the dataset imputed using the Vtreat methodology. The tree based nature of this algorithm allowed us to export the relative importance or contribution of each input variable in predicting the response.

Let *T*, be a single decision tree, based on [60]. We apply the following equation as an index of relevance for each of our features (predictor variables) *X*_*R*_:

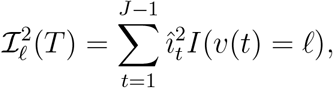

where *J* − 1 indicates the internal nodes of the tree, *t*, indicates each one of the input variables *X*_*v*(*t*)_ and 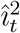 indicates the maximal estimated improvement which defines the particular variable in squared error risk over that for a constant fit over the entire region 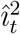. Utilizing this equation through the XGBoost model [47], we obtain a list with the importance of each feature.

The outcomes illustrated in Figure 3 offer new insights for the HealthStatus evaluation. More specifically, the most important variables (left plot in Figure 3) regarding their effectiveness in the HealthStatus prediction were the features: “Respondent’s self-rated/self-reported health” (srh), “Age at time of measure” (age), “History of arthritis, rheumatism or osteoarthritis” (“h_joint_disorders”), “Current depressive status” (depression), “Engage in vigorous exercise during the last two weeks” (vig_pa), “Highest score, to the nearest kg, of all hand grip measurements, regardless” (grip), “Psychological measure of anxiety symptoms” (anxiety_symp), “Weekly frequency of less vigorous exercise” (f_mod_pa), “Participant has paid employment” (employed), and “Highest level of formal education achieved” (education).

**Figure 3:**
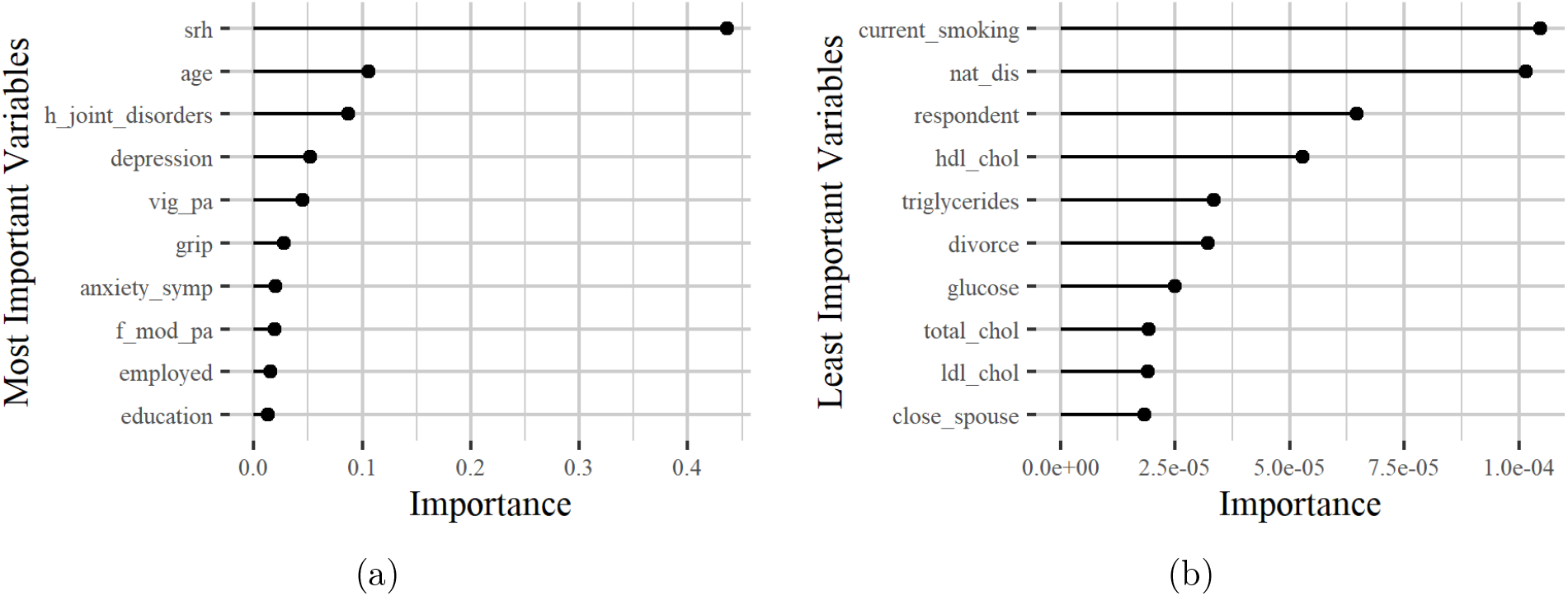
The horizontal bars illustrate the most (left) and the least (right) important variables regarding their effectiveness in the HealthStatus prediction by applying the XGBoost classification algorithm. The x-axis imprints the variable importance score while the y-axis includes the feature names defined by the ATHLOS project (see supplementary sheet S1).

On the other hand, the least important features (right plot in Figure 3) were the “Current smoker, any type of tobacco smoking” (current_smoking), “Ever experienced any natural disaster” (nat_dis), “Respondent is the participant, spouse/partner or other” (respondent), “Serum HDL Cholesterol” (hdl_chol), “Triglycerides” (triglycerides), “Experience of divorce/separation” (divorce), “Blood/serum glucose levels” (glucose), “Total Cholesterol” (total_chol), “Serum LDL Cholesterol” (ldl_chol) and “Is the relationship with the spouse close” (close_spouse).

Through the variable importance analysis we interestingly observe that there are factors that significantly affect HealthStatus prediction, while being unrelated to health, as well as factors that do not affect the HealthStatus but apparently should have, based on the recent literature. More specifically, the levels of depression were considered an important factor that affects the respondent’s HealthStatus prediction, a condition that exists at various age levels and does not necessarily indicate aging. Factors such as Serum HDL Cholesterol, Triglycerides, Blood/serum glucose levels, and Total Cholesterol were considered minor to determine the accurate HealthStatus prediction. However, all these factors are quite relevant to the human status health [61, 62].

Additionally, it is impressive how relevant is everyone’s personal opinion about assessing their state of health. Our results indicated that it is the most important factor. We may consider encouraging the fact that most can personally assess their state of health, since this could affect prevention of adverse events and bad incidents, contributing to the Medical and Healthcare domain’s effectiveness that follows the participatory and preventive factors, two of the four main axes that constitute the future of Medicine [63].

## 4 Concluding Remarks

In this paper, we emphasize on the development of a complete ML methodology for the ATHLOS (Ageing Trajectories of Health: Longitudinal Opportunities and Synergies) Project, aiming in the accurate prediction of the HealthStatus (HS) score, an index that estimates the human status of health. We deal with the most critical aspect of the inherent complexity of the provided dataset, the high degree of missing values, by exploring extensively and discovering the most effective methodologies for missing value imputation. Our results indicated the effect of different imputation strategies on Machine Learning tasks exposing the capacity of accurate predictions when combined with cutting-edge tools. We additionally concluded that the highlighted Vtreat imputation method can be rendered as an imputation tool-guide for multiple unified, independent longitudinal studies. Finally, the variable importance analysis highlighted new factors that may affect the state of human health exposing the potential of further findings within similar large scale longitudinal studies. In our future research, we intend to further investigate the development of a unified model on the basis of a bidirectional ML scheme for data imputation and prediction.

## Data Availability

The data contained in this paper are not publicly available, they can be obtained from through the ATHLOS project.

## Acknowledgement

This work is supported by the ATHLOS (Ageing Trajectories of Health: Longitudinal Opportunities and Synergies) project, funded by the European Union’s Horizon 2020 Research and Innovation Programme under grant agreement number 635316.

